# Continuous Assessment of Daily-Living Gait Using Self-Supervised Learning of Wrist-Worn Accelerometer Data

**DOI:** 10.1101/2025.05.21.25328061

**Authors:** Yonatan E. Brand, Aron S. Buchman, Felix Kluge, Luca Palmerini, Clemens Becker, Andrea Cereatti, Walter Maetzler, Beatrix Vereijken, Alison J. Yarnall, Lynn Rochester, Silvia Del Din, Arne Mueller, Jeffrey M. Hausdorff, Or Perlman

**Author notes:** Correspondence: Or Perlman.

## Abstract

Physical activity and mobility are critical for healthy aging and predict diverse health outcomes. While wrist-worn accelerometers are widely used to monitor physical activity, estimating gait metrics from wrist data remains challenging. We extend ElderNet, a self-supervised deep-learning model previously validated for walking-bout detection, to estimate gait metrics from wrist accelerometry. Validation involved 819 older adults (Rush-Memory-and-Aging-Project) and 85 individuals with gait impairments (Mobilise-D), from six medical centers. In Mobilise-D, ElderNet achieved an absolute error of 8.82 cm/s and an intra-class correlation of 0.87 for gait speed, outperforming state-of-the-art methods (p < 0.001) and models using a lower-back sensor. ElderNet outperformed (percentage error; p < 0.01) competing approaches in estimating cadence and stride length, and better (p < 0.01) classified mobility disability (AUC = 0.80) than conventional gait or physical activity metrics. These results render ElderNet a scalable tool for gait assessment using wrist-worn devices in aging and clinical populations.

## INTRODUCTION

Physical activity (PA) and mobility are essential components of healthy aging and are strong predictors of a wide range of clinical outcomes, including cognitive decline, disability, and mortality^1–6^. Objective measures of PA—such as sedentary time and moderate-to-vigorous PA— are conventionally measured using wrist-worn accelerometers and offer valuable insights^7,8^. However, these measures typically capture overall physical activity levels and fail to distinguish between specific types of physical activity, such as gait versus other daily movements, limiting the ability to identify the distinct health benefits associated with different forms of activity^9^. Gait-specific measures, such as gait speed, cadence, and stride length, have been shown to serve as potential biomarkers, potentially providing complimentary insight and objective measures of aging, functional health, and disease progression^10–13^. Thus, untangling gait from total daily physical activity may enable more precise identification of activity types to optimize the prediction of health outcomes in older populations and inform targeted interventions.

Traditionally, the assessment of gait quality has been conducted under controlled laboratory or clinical conditions, often using short walking tests or instrumented walkways^14,15^. While these methods offer high accuracy, they only reflect brief snapshots of an individual’s mobility and may miss important variations that occur during daily life^16–18^. Recent advancements in wearable sensor technology have opened new opportunities to monitor gait in free-living, daily-life conditions, enabling long-term, remote assessment in natural environments^19,20^. Most real-world gait studies to date have relied on accelerometers placed on the lower back, trunk, legs or feet^14,21,22^. From a signal processing perspective, these sensor locations benefit from proximity to the body’s center of mass or ground contact. These sensor placements allow for accurate spatio-temporal gait estimates^14^, but comfort and usability concerns may limit long-term adherence and scalability^23^.

In contrast, wrist-worn accelerometers are widely adopted in both clinical and consumer settings due to their convenience and high user compliance^8,15,24^. Large-scale population studies, such as the UK Biobank, have successfully demonstrated the feasibility of using wrist-worn accelerometers to derive meaningful population-level physical activity metrics, including step counts^5,8,25–27^. For example, Doherty et al. developed open-source algorithms for processing wrist accelerometry data, facilitating foundational insights into physical activity patterns across more than 100,000 participants^5,28–31^. However, these efforts have predominantly focused on overall physical activity quantification rather than on detailed assessments of spa-tial-temporal measures of gait, highlighting a critical gap and untapped potential.

The wrist’s distance from the body’s center of mass and the complexity of its motion during walking pose significant challenges for extracting meaningful and accurate gait metrics^32^. As a result, only a limited number of studies have attempted to estimate the spatial-temporal measures of gait from wrist data in real-world conditions. The few that do have largely focused on healthy individuals, rather than older adults or those with gait impairment^15,33^, and often necessitate individual-level calibration^34,35^.

Multi-day wrist recordings capture all of the movements of arm and hand. These recordings can include varied activities of the arm as well as walking. Two steps are necessary to extract meaningful gait metrics from wrist data. First, tools are needed to differentiate bouts of gait from non-gait movements throughout the entire recording. Once the bouts of gait are identified, additional tools are needed to quantify conventional gait characteristics, including speed, cadence and stride length, and regularity. In a recent study, we introduced and validated a self-supervised deep-learning framework, ElderNet, that accurately identifies bouts of gait from 3D wrist data^20^. ElderNet leverages a self-supervised learning (SSL) approach, utilizing large amounts of unlabeled accelerometry data to learn robust, generalizable representations of gait patterns. The framework comprises two main steps: an initial SSL-based pretraining using the large unlabeled UK Biobank dataset^5,8,25^ (Dataset 1) followed by additional SSL training on the RUSH Memory and Aging Project (MAP) dataset^36,37^ (Dataset 2) tailored toward older populations with diverse mobility characteristics and impairments. The second step involves fine-tuning the foundational SSL model for a specific downstream task, such as gait speed estimation, using a smaller, labeled dataset. Here, we adapted and expanded ElderNet to estimate detailed gait metrics for each identified walking bout, building on its previously demonstrated ability to detect walking segments from wrist-worn accelerometry.

More specifically, we fine-tuned ElderNet using a labeled dataset from the Mobilise-D technical validation study (TVS; Dataset 3), a relatively large, daily living, labeled dataset comprising approximately 2.5-hours recordings from older adults and individuals with gait impairments, alongside robust reference annotations obtained from a validated multi-sensor reference system that included pressure insoles^38^. This dataset provides extensive real-world annotations of spatio-temporal gait parameters, enabling accurate model training and validation^39,40^. ElderNet’s performance was evaluated on the test set of the TVS dataset (Dataset 3) against a fully supervised baseline model with the same architecture as ElderNet (but without SSL pretraining) to quantify the benefit of the SSL component. For gait speed estimation, we also compared ElderNet to a state-of-the-art biomechanical approach proposed by Soltani et al.^33^, adapted to our dataset. In addition, to further assess generalizability, we validated ElderNet on an independent dataset of 11 healthy young adults (Dataset 4). Finally, to demonstrate ElderNet’s potential clinical utility in real-world contexts, we applied it to a multi-day, free-living data from the RUSH MAP^36,37^ (Dataset 2). Specifically, we examined whether the multi-day gait quality metrics extracted by ElderNet could effectively classify mobility disability status in older adults. We also directly compared the discriminative performance of multi-day gait metrics derived from ElderNet to metrics obtained from traditional supervised short assessments of gait and mobility, thereby demonstrating the added value of continuous real-world monitoring. Furthermore, we compared the model based on multi-day gait metrics with a model using total daily physical activity metrics extracted from the same recordings to explore the added value of detailed multi-day gait metrics.

## RESULTS

### Evaluation of ElderNet for Gait Speed Estimation

To evaluate ElderNet’s ability to estimate real-world gait quality metrics, we fine-tuned the model using the labeled dataset from the Mobilise-D TVS (Dataset 3). Figure 1 illustrates the ElderNet pipeline, and Table 1 summarizes the key characteristics of the Mobilise-D TVS dataset. Further details, including dataset composition and training protocols, are provided in Table 2 and the Methods section.

**Figure 1.**
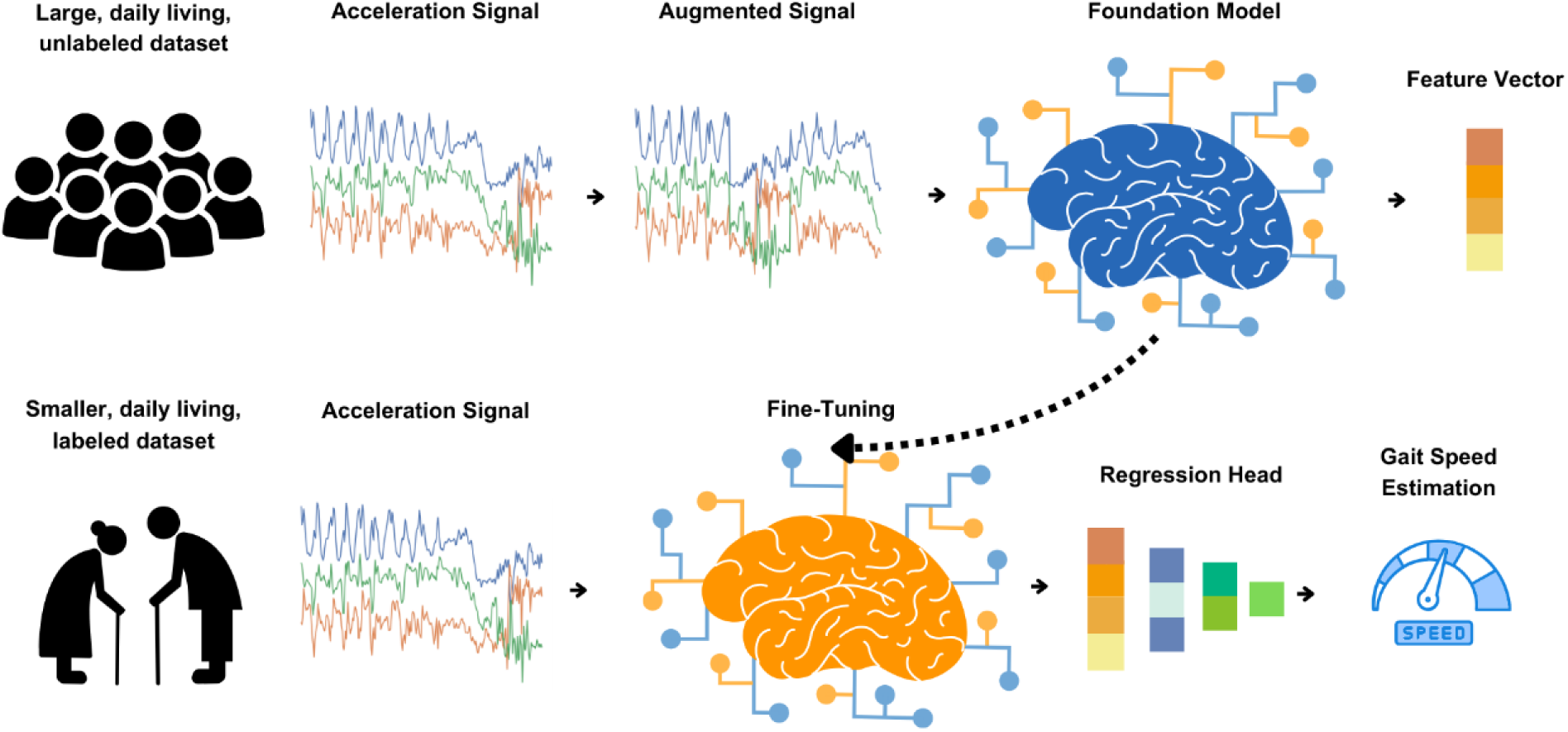
ElderNet Pipeline. First, an SSL model pre-trained on the UK Biobank dataset was adapted for older adults by adding trainable layers and using data from the RUSH MAP. This adapted model, ElderNet, served as the foundation model. In the second stage, ElderNet was fine-tuned for gait quality estimation using labeled data from the Mobilise-D dataset, with a regression head added to extract gait-related features from acceleration signals.

**Table 1.**
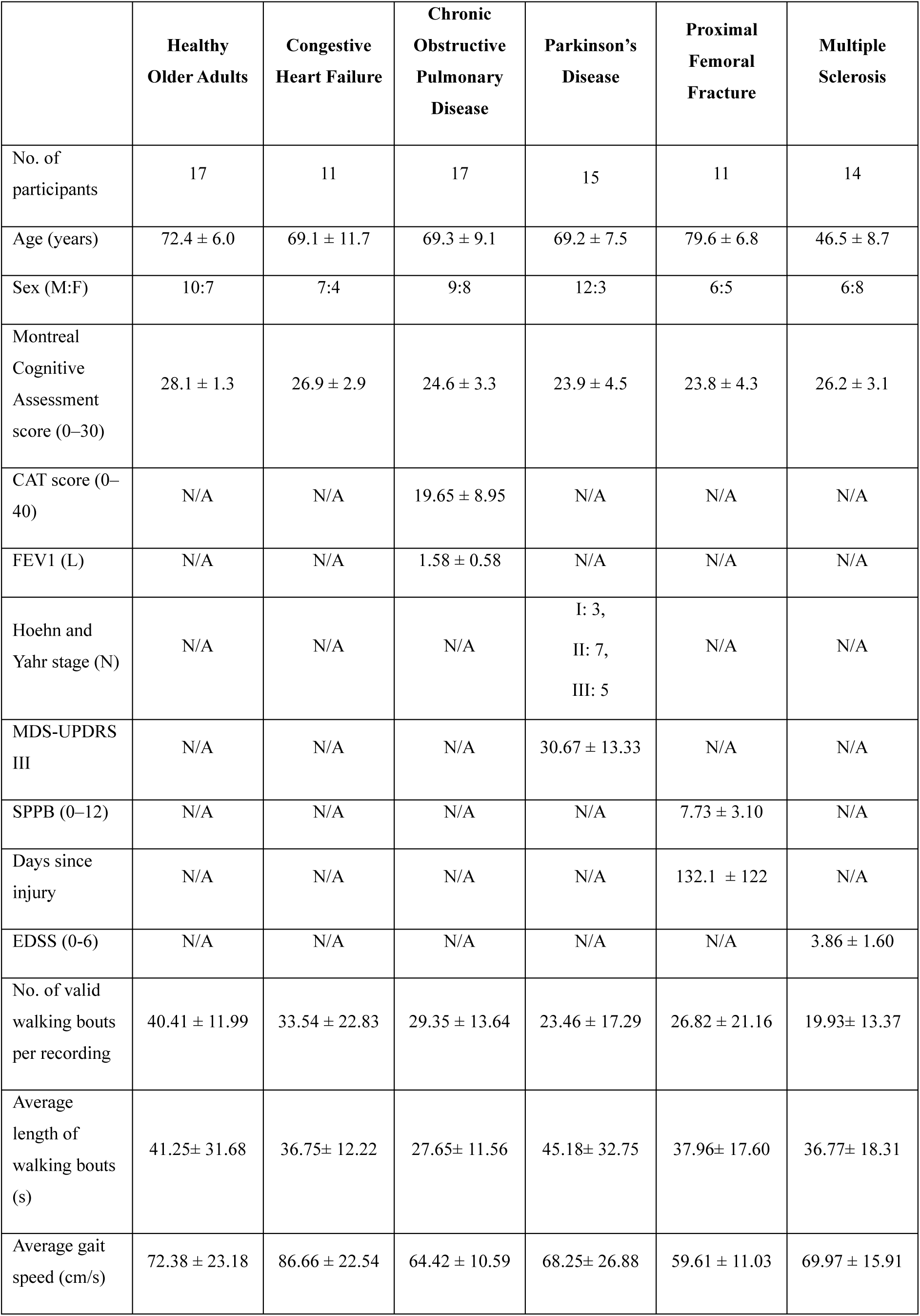
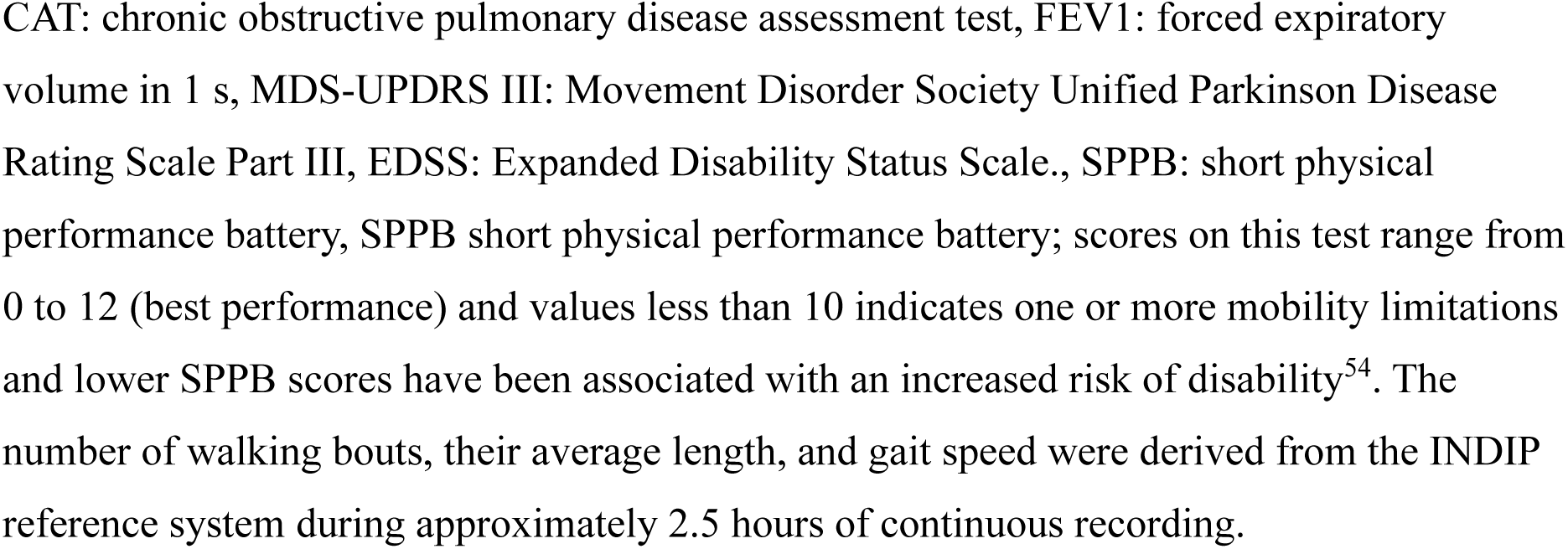
Characteristics of older adults in the Mobilise-D technical validation study (Dataset 3)

|  | **Healthy**<br>**Older Adults** | **Congestive**<br>**Heart Failure** | **Chronic**<br>**Obstructive**<br>**Pulmonary**<br>**Disease** | **Parkinson's**<br>**Disease** | **Proximal**<br>**Femoral**<br>**Fracture** | **Multiple**<br>**Sclerosis** |
| --- | --- | --- | --- | --- | --- | --- |
| No. of participants | 17 | 11 | 17 | 15 | 11 | 14 |
| Age (years) | 72.4 ± 6.0 | 69.1 ± 11.7 | 69.3 ± 9.1 | 69.2 ± 7.5 | 79.6 ± 6.8 | 46.5 ± 8.7 |
| Sex (M:F) | 10:7 | 7:4 | 9:8 | 12:3 | 6:5 | 6:8 |
| Montreal Cognitive Assessment score (0–30) | 28.1 ± 1.3 | 26.9 ± 2.9 | 24.6 ± 3.3 | 23.9 ± 4.5 | 23.8 ± 4.3 | 26.2 ± 3.1 |
| CAT score (0–40) | N/A | N/A | 19.65 ± 8.95 | N/A | N/A | N/A |
| FEV1 (L) | N/A | N/A | 1.58 ± 0.58 | N/A | N/A | N/A |
| Hoehn and Yahr stage (N) | N/A | N/A | N/A | I: 3,<br>II: 7,<br>III: 5 | N/A | N/A |
| MDS-UPDRS III | N/A | N/A | N/A | 30.67 ± 13.33 | N/A | N/A |
| SPPB (0–12) | N/A | N/A | N/A | N/A | 7.73 ± 3.10 | N/A |
| Days since injury | N/A | N/A | N/A | N/A | 132.1 ± 122 | N/A |
| EDSS (0-6) | N/A | N/A | N/A | N/A | N/A | 3.86 ± 1.60 |
| No. of valid walking bouts per recording | 40.41 ± 11.99 | 33.54 ± 22.83 | 29.35 ± 13.64 | 23.46 ± 17.29 | 26.82 ± 21.16 | 19.93± 13.37 |
| Average length of walking bouts (s) | 41.25± 31.68 | 36.75± 12.22 | 27.65± 11.56 | 45.18± 32.75 | 37.96± 17.60 | 36.77± 18.31 |
| Average gait speed (cm/s) | 72.38 ± 23.18 | 86.66 ± 22.54 | 64.42 ± 10.59 | 68.25± 26.88 | 59.61 ± 11.03 | 69.97 ± 15.91 |

CAT: chronic obstructive pulmonary disease assessment test, FEV1: forced expiratory volume in 1 s, MDS-UPDRS III: Movement Disorder Society Unified Parkinson Disease Rating Scale Part III, EDSS: Expanded Disability Status Scale., SPPB: short physical performance battery, SPPB short physical performance battery; scores on this test range from 0 to 12 (best performance) and values less than 10 indicates one or more mobility limitations and lower SPPB scores have been associated with an increased risk of disability<sup>54</sup>. The number of walking bouts, their average length, and gait speed were derived from the INDIP reference system during approximately 2.5 hours of continuous recording.

**Table 2.** Summary of datasets used in this study.

| **Dataset #** | **Name** | **Sample Size** | **Population Characteristics** | **Duration** | **Purpose in Study** | **Annotation type** |
| --- | --- | --- | --- | --- | --- | --- |
| Dataset 1 | UK Biobank | >100,00 | General population | 7 days | SSL pretraining | Unlabeled |
| Dataset 2 | RUSH MAP | 819 | Older adults | 10 days | SSL adaptation; Clinical validation | Mobility disability scores |
| Dataset 3 | Mobilise-D TVS | 85 | Older adults & gait impaired | 2.5 hours | Fine-tuning & evaluation | Spatio-temporal gait annotations |
| Dataset 4 | Healthy young adults (Mobilise-D) | 11 | Healthy young adults | 2.5 hours | External validation | Spatio-temporal gait annotations |

All reported results reflect performance on the Mobilise-D TVS test set (20% of dataset 3), which includes representative individuals from each clinical cohort. On this test set, ElderNet demonstrated high performance in gait speed estimation, achieving median values across subjects for mean absolute error (MAE) of 8.8 cm/s, root mean square error (RMSE) of 10.7 cm/s, mean absolute percentage error (MAPE) of 12.73%, coefficient of determination (R²) of 0.74, and, compared to measures obtained from the gold-standard pressure sensitive insole system, an intra-class correlation coefficient ICC(2,1) of 0.87. As shown in Figure 2, ElderNet significantly outperformed both the current state-of-the-art model by Soltani et al.^33^ and a fully supervised baseline model, which uses the same architecture as ElderNet but is trained from scratch in a purely supervised manner, without any self-supervised pretraining phase. Furthermore, ElderNet maintained its superior performance on an external dataset (Dataset 4) comprising healthy young adults, achieving an MAE of 8.3 cm/s, RMSE of 11.4 cm/s, MAPE of 8.22%, R² of 0.72, and ICC of 0.87, and significantly outperforming the supervised baseline in both RMSE (p<0.05) and ICC (p<0.01). Detailed performance metrics, including the 25th and 75th percentiles at the subject level, are provided in Table S1. Figure 3 further illustrates these findings, showing a stronger correlation between ElderNet’s predictions and true gait speeds, compared to alternative models.

**Figure 2.**
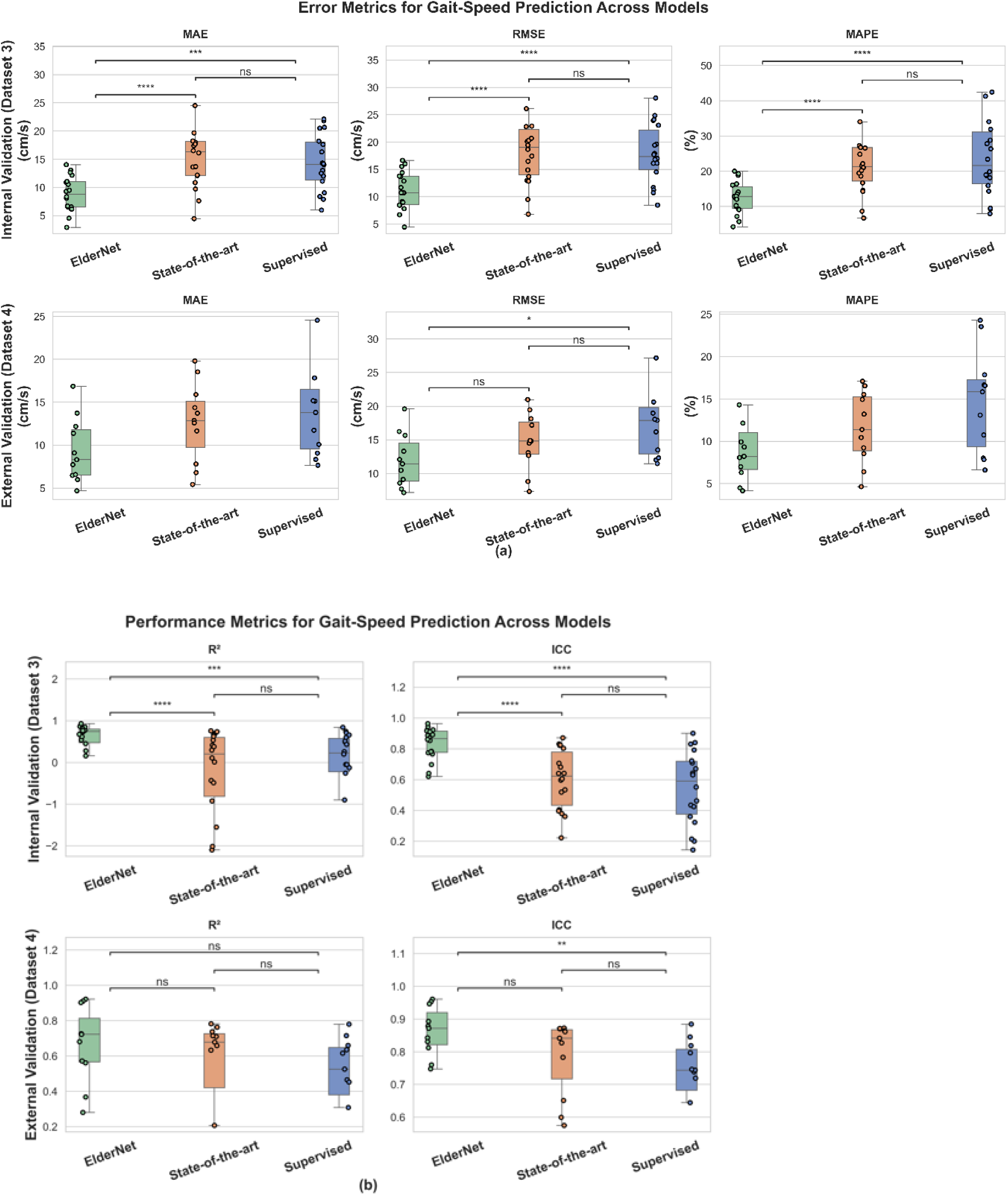
Performance comparison of ElderNet with benchmark models for gait-speed prediction. Boxplots show subject-level error (MAE, RMSE, MAPE) and performance (R², ICC) for ElderNet, state-of-the-art, and supervised baseline models on the internal Mobilize-D TVS dataset (Dataset 3; n =18) and external validation with healthy young adults (Dataset 4; n=11). Outliers were excluded from plots for clarity but included in statistical analyses. Overall model differences were assessed using a Friedman test (p < 0.05); significant results were followed by Wilcoxon signed-rank tests for pairwise comparisons. Asterisks denote significance: *p < 0.05; **p < 0.01; ***p < 0.001; ****p < 0.0001; ns = not significant.

**Figure 3.**
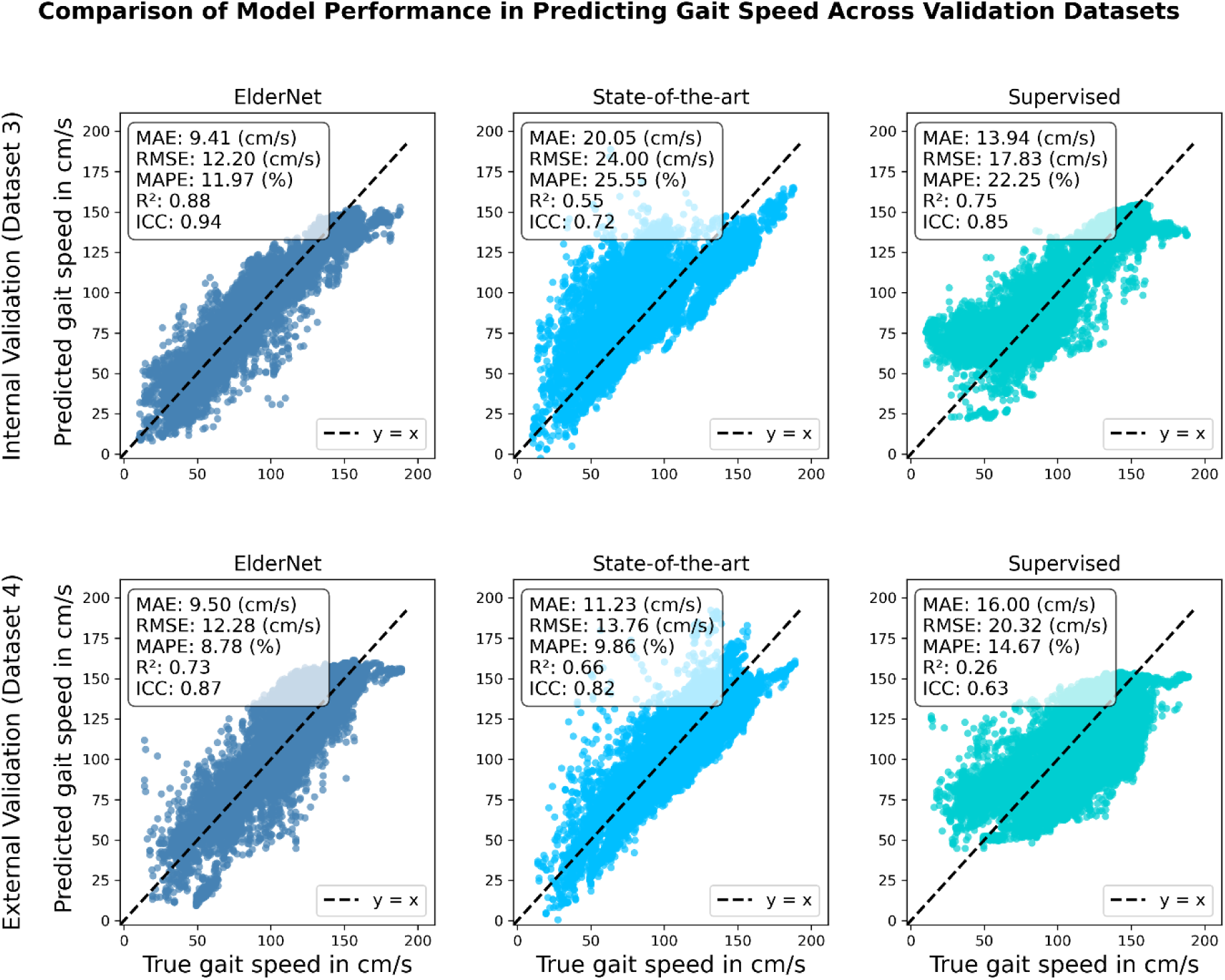
Correlation plots comparing the proposed ElderNet model with state-of-the-art and supervised baseline models. The scatter plots depict predicted versus true gait speed for older adults and individuals with gait impairments (Dataset 3 test set) and young adults external dataset (Dataset 4), with each point representing a 10-second window. The error and performance metrics are at the window level as well. ElderNet demonstrated superior predictive performance across both cohorts. The dashed line (y = x) indicates perfect agreement between predicted and true values.

The Bland-Altman plot in Figure 4 compares ElderNet’s gait speed predictions with reference measurements from various Mobilise-D TVS cohorts. This analysis revealed a minimal mean error of 0.17 cm/s, indicating low prediction bias, notably lower than a lower-back-based algorithm tested on the same cohorts^23^. Table 3 provides a summary of ElderNet’s gait speed estimation performance across the different cohorts in the Mobilise-D TVS dataset.

**Figure 4.**
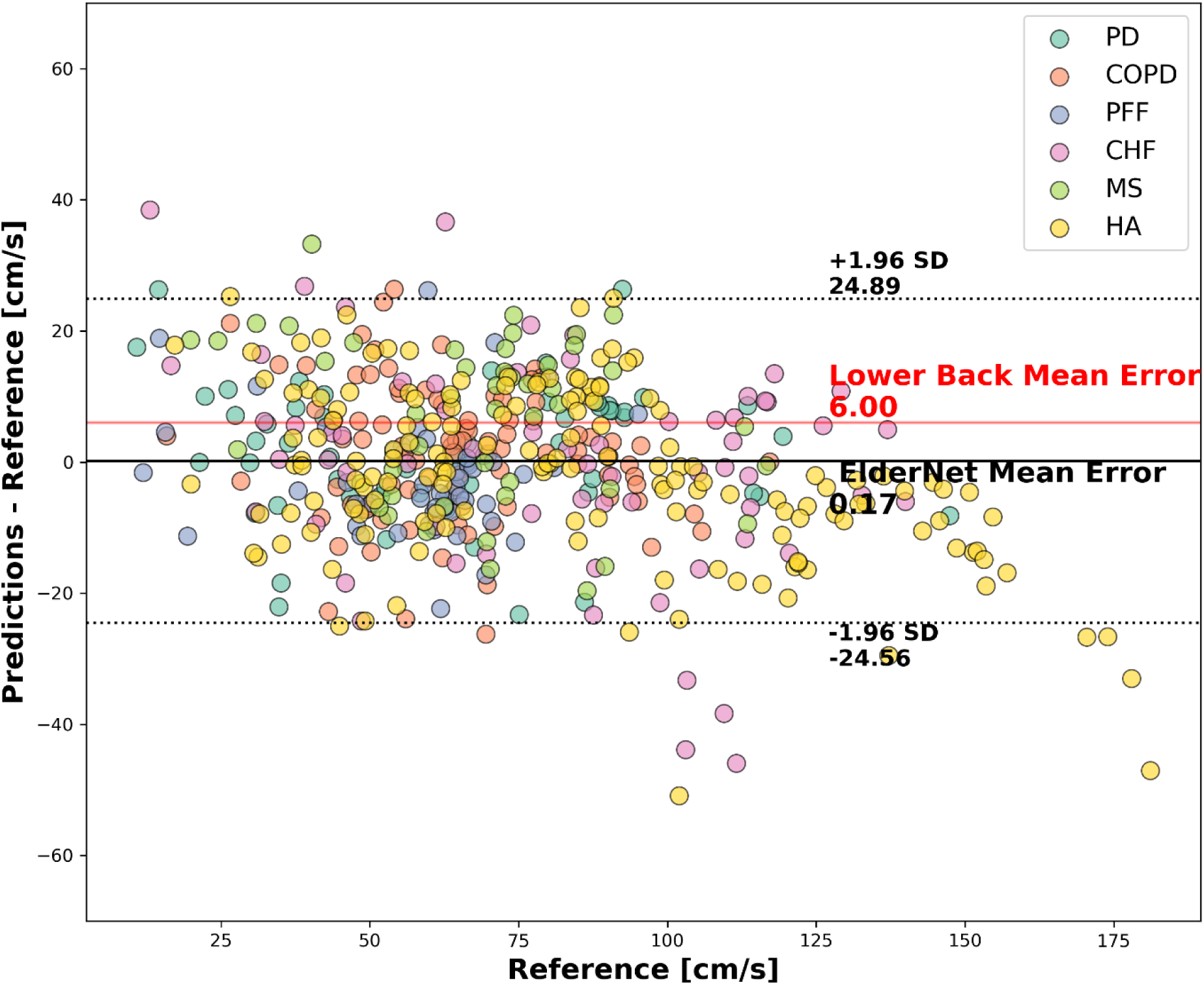
Bland-Altman plot comparing gait speed predictions to ground truth in the Mobilise-D TVS test set (Dataset 3; n=18). Each dot represents a walking bout with a minimum duration of 10 seconds. The horizontal dashed lines indicate the 95% confidence intervals around the mean error. The red line represents the mean error of a lower-back-based model evaluated on the entire Mobilise-D dataset (n=82) ^23^.

**Table 3.**
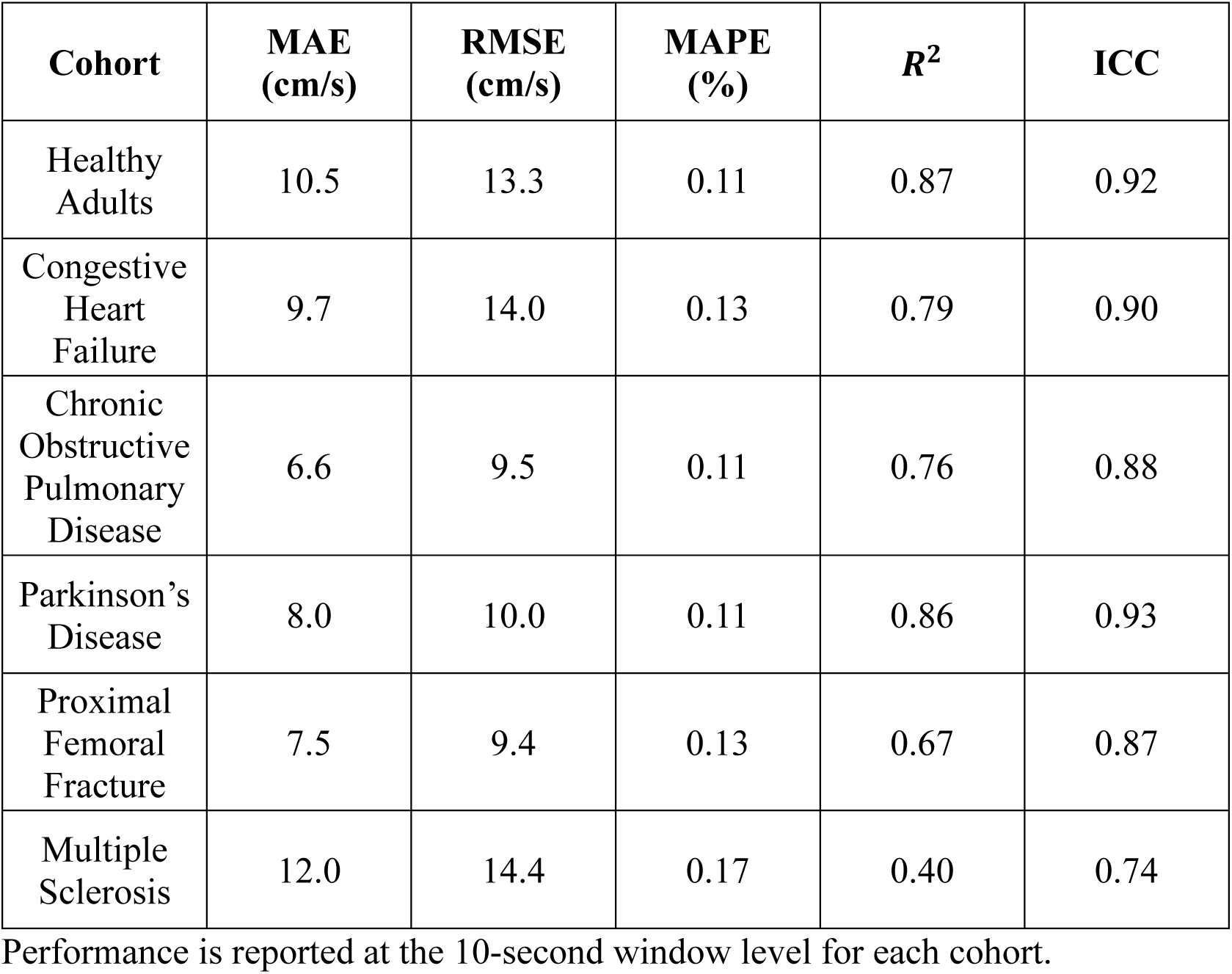
ElderNet performance for gait speed across different cohorts in the Mobilise-D TVS (Dataset 3) test set (n=18)

### Evaluation of ElderNet for additional Gait Metrics

Beyond gait speed estimation, ElderNet was extended to quantify additional gait metrics, including cadence, stride length, and stride regularity, to provide a more comprehensive assessment of gait quality during daily living (Table 4). For cadence, ElderNet achieved a median MAE of 3.11 steps/min, RMSE of 5.19 steps/min, MAPE of 3.71%, R² of 0.65, and an ICC of 0.83. Compared with a fully supervised model, ElderNet showed significantly lower errors (MAE: p < 0.001; RMSE: p = 0.045), higher ICC (p = 0.02), and a trend towards a higher R², though this did not reach statistical significance (p = 0.07).

**Table 4.** Comparison of ElderNet and a supervised model for estimating gait quality metrics in the Mobilise-D TVS (Dataset 3) test set.

| Measure | Unit | Facet of Mobility | Metric | ElderNet<br>(median<br>[25<sup>th</sup>-75<sup>th</sup>]) | Supervised<br>(median<br>[25<sup>th</sup>-75<sup>th</sup>]) | p-value |
| --- | --- | --- | --- | --- | --- | --- |
| **Cadence** | steps/minute | Rhythm | MAE | 3.11<br>[2.26 3.88] | 3.77<br>[2.94 4.46] | **<0.001\*\*\*** |
|  |  |  | RMSE | 5.19<br>[3.79 6.48] | 5.83<br>[4.72 6.88] | **0.045\*** |
|  |  |  | MAPE | 3.71<br>[2.54 4.54] | 4.26<br>[3.27 5.39] | **<0.01\*\*** |
|  |  |  | R<sup>2</sup> | 0.65<br>[0.58 0.74] | 0.55<br>[0.45 0.74] | 0.07 |
|  |  |  | ICC | 0.83<br>[0.75 0.87] | 0.77<br>[0.69 0.86] | **0.02\*** |
| **Stride Length** | cm | Pace | MAE | 11.61<br>[8.83 13.68] | 14.57<br>[10.90 17.43] | **<0.01\*\*** |
|  |  |  | RMSE | 14.09<br>[11.13 16.59] | 18.30<br>[14.47 20.64] | **<0.001\*\*\*** |
|  |  |  | MAPE | 11.09<br>[8.43 14.39] | 15.62<br>[12.56 21.54] | **<0.001\*\*\*** |
|  |  |  | R<sup>2</sup> | 0.48<br>[0.14 0.69] | 0.31<br>[-0.62 0.45] | **<0.001\*\*\*** |
|  |  |  | ICC | 0.72<br>[0.59 0.83] | 0.54<br>[0.35 0.64] | **<0.001\*\*\*** |
| **Stride Regularity** | A.U. (0-1) | Variability | MAE | 0.08<br>[0.08 0.10] | 0.08<br>[0.07 0.09] | 0.30 |
|  |  |  | RMSE | 0.11<br>[0.10 0.13] | 0.11<br>[0.09 0.12] | 0.21 |
|  |  |  | MAPE | 28.25<br>[23.25 36.36] | 31.27<br>[19.45 35.82] | 0.30 |
|  |  |  | R<sup>2</sup> | 0.55<br>[0.38 0.69] | 0.67<br>[0.42 0.76] | 0.30 |
|  |  |  | ICC | 0.76<br>[0.68 0.82] | 0.84<br>[0.75 0.88] | **0.012** |

For stride length, ElderNet also demonstrated superior performance (MAE: 10.6 cm, p < 0.01; RMSE: 13.1, MAPE: 11.09%, R²: 0.53, ICC: 0.78; p < 0.001 for all others) compared with the supervised approach. Stride regularity estimation by ElderNet exhibited performance comparable to the supervised model, with both models achieving similar error rates (MAE = 0.08 A.U., RMSE = 0.11 A.U.; A.U. = arbitrary unit), though the supervised model indicated a slightly higher ICC (0.84 vs. 0.76) and R² (0.67 vs. 0.55). However, none of these differences were statistically significant (all p-values > 0.05).

### Demonstration of Potential Clinical Utility

As a preliminary demonstration of ElderNet’s potential clinical utility, we applied it to continuous, ∼10-day wrist accelerometer recordings from the RUSH MAP dataset (Dataset 2), extracting daily-living gait quality measures (see Methods). Participant characteristics are summarized in Table 5. A heat map summarizing the correlations between measures obtained using the two approaches (Figure S1) revealed that gait characteristics recorded in supervised assessments are only mildly related to the same gait characteristics extracted from daily living conditions. Although internal consistency within settings was strong (supervised lower-back features correlated well among themselves, and similarly for wrist-based daily-living features), cross-setting correlations were weaker, suggesting that supervised gait measures do not fully reflect daily-living gait behavior.

**Table 5.**
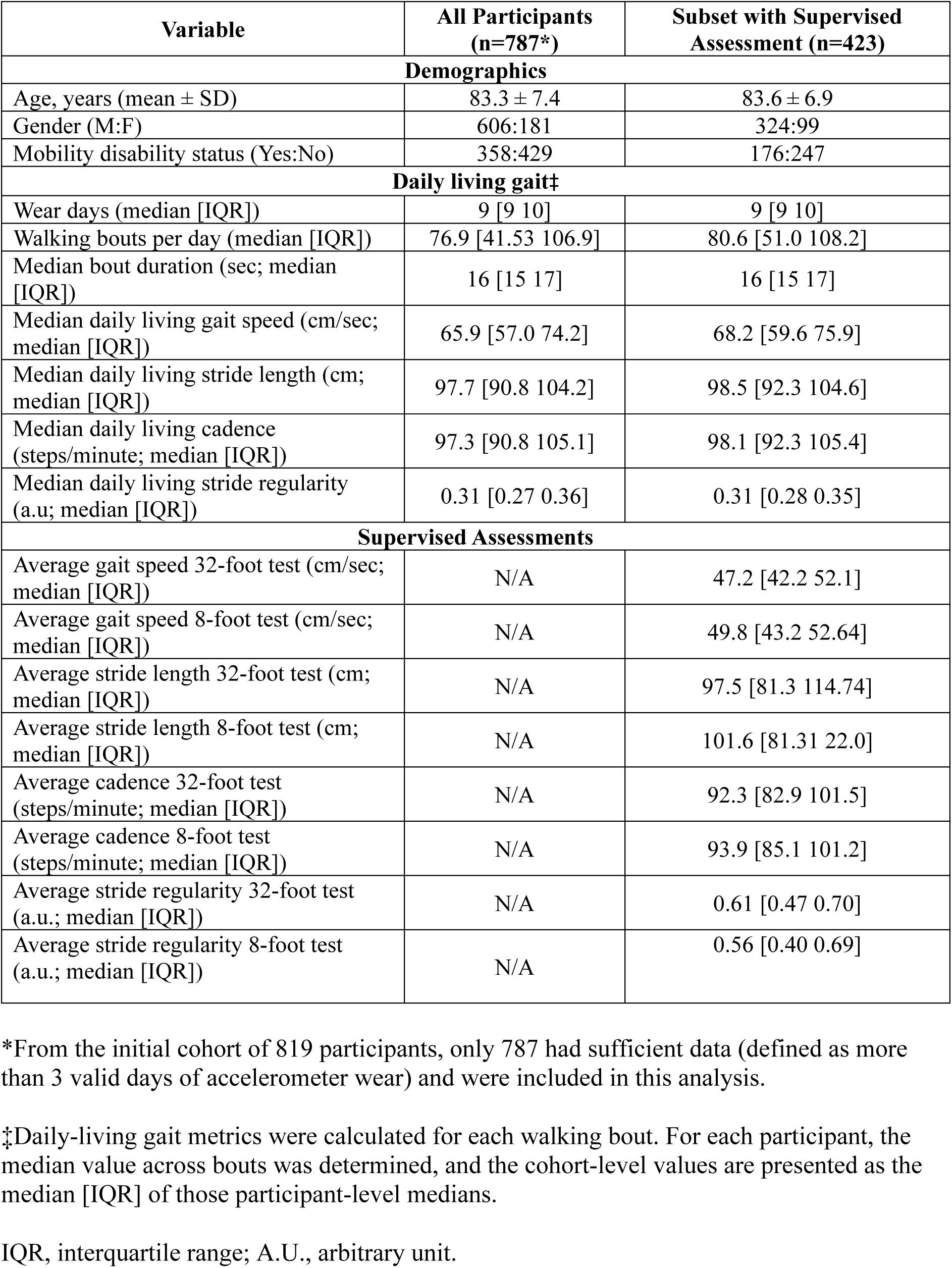
Characteristics of the RUSH Memory and Aging project cohort (Dataset 2)

| Variable | All Participants<br>(n=787*) | Subset with Supervised<br>Assessment (n=423) |
| --- | --- | --- |
| **Demographics** |  |  |
| Age, years (mean ± SD) | 83.3 ± 7.4 | 83.6 ± 6.9 |
| Gender (M:F) | 606:181 | 324:99 |
| Mobility disability status (Yes:No) | 358:429 | 176:247 |
| **Daily living gait‡** |  |  |
| Wear days (median [IQR]) | 9 [9 10] | 9 [9 10] |
| Walking bouts per day (median [IQR]) | 76.9 [41.53 106.9] | 80.6 [51.0 108.2] |
| Median bout duration (sec; median [IQR]) | 16 [15 17] | 16 [15 17] |
| Median daily living gait speed (cm/sec; median [IQR]) | 65.9 [57.0 74.2] | 68.2 [59.6 75.9] |
| Median daily living stride length (cm; median [IQR]) | 97.7 [90.8 104.2] | 98.5 [92.3 104.6] |
| Median daily living cadence (steps/minute; median [IQR]) | 97.3 [90.8 105.1] | 98.1 [92.3 105.4] |
| Median daily living stride regularity (a.u; median [IQR]) | 0.31 [0.27 0.36] | 0.31 [0.28 0.35] |
| **Supervised Assessments** |  |  |
| Average gait speed 32-foot test (cm/sec; median [IQR]) | N/A | 47.2 [42.2 52.1] |
| Average gait speed 8-foot test (cm/sec; median [IQR]) | N/A | 49.8 [43.2 52.64] |
| Average stride length 32-foot test (cm; median [IQR]) | N/A | 97.5 [81.3 114.74] |
| Average stride length 8-foot test (cm; median [IQR]) | N/A | 101.6 [81.31 22.0] |
| Average cadence 32-foot test (steps/minute; median [IQR]) | N/A | 92.3 [82.9 101.5] |
| Average cadence 8-foot test (steps/minute; median [IQR]) | N/A | 93.9 [85.1 101.2] |
| Average stride regularity 32-foot test (a.u.; median [IQR]) | N/A | 0.61 [0.47 0.70] |
| Average stride regularity 8-foot test (a.u.; median [IQR]) | N/A | 0.56 [0.40 0.69] |

\*From the initial cohort of 819 participants, only 787 had sufficient data (defined as more than 3 valid days of accelerometer wear) and were included in this analysis.
‡Daily-living gait metrics were calculated for each walking bout. For each participant, the median value across bouts was determined, and the cohort-level values are presented as the median [IQR] of those participant-level medians.
IQR, interquartile range; A.U., arbitrary unit.

To evaluate the discriminative utility of gait metrics collected in daily life, we compared performance against gait metrics obtained in a controlled, supervised setting, as well as PA features derived from daily living wrist accelerometry. We trained an XGBoost classifier to classify mobility disability status (defined using the Rosow-Breslau scale). As illustrated in Figure 5, the supervised-based model achieved an area under the curve (AUC) of 0.67 ± 0.09. In comparison, the model using PA features from daily-living wrist accelerometry performed slightly better (AUC: 0.69 ± 0.06). The highest accuracy was achieved by a model incorporating the daily life gait metrics (AUC: 0.80 ± 0.06).

**Figure 5.**
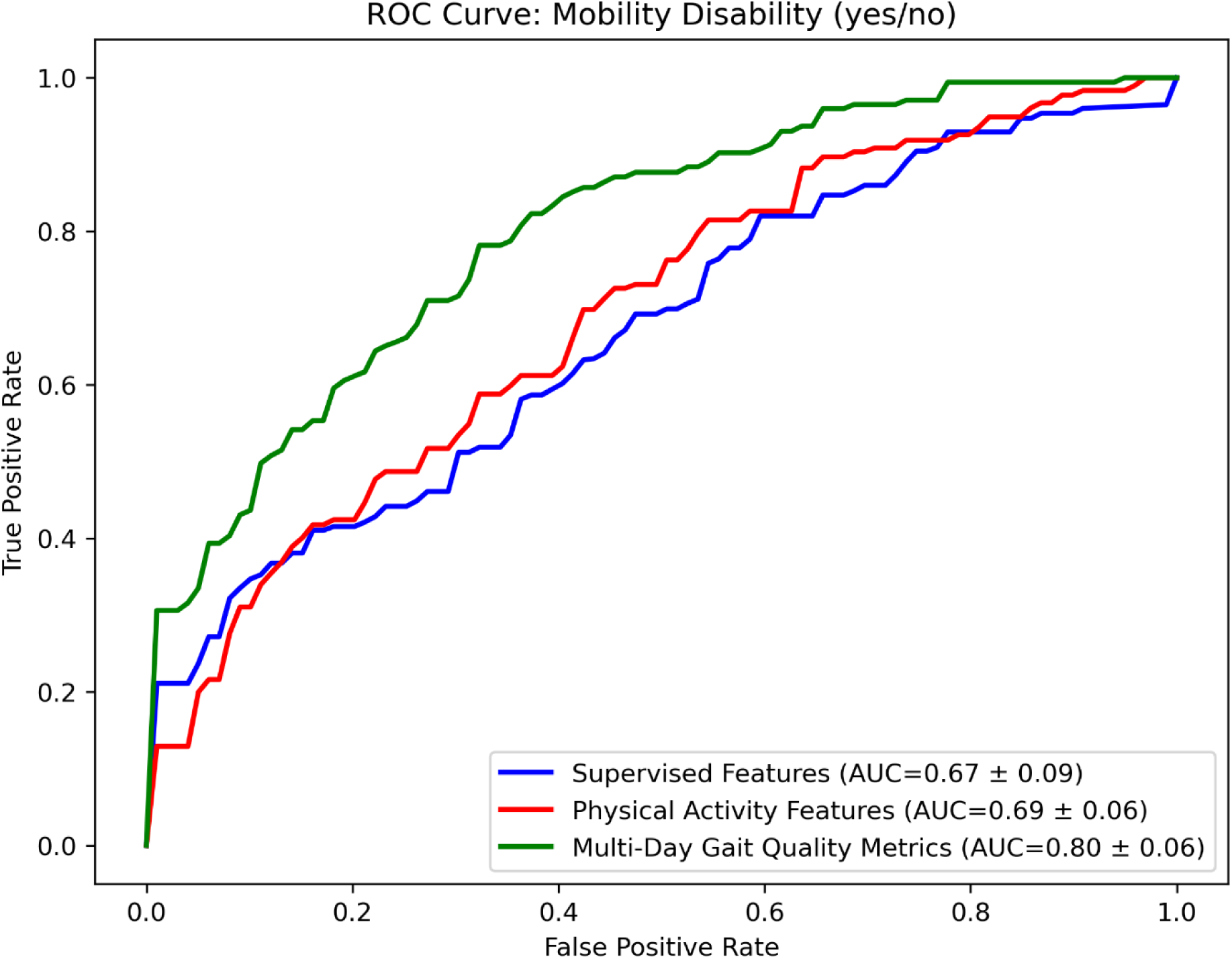
ROC Curve for classifying mobility disability in the RUSH Memory and Aging project (Dataset 2; n = 423). Curves represent mean values (±1 SD across 10 folds) obtained from nested cross-validation. Three different feature sets were compared: supervised-based accelerometry features (blue) extracted from standardized walking tests (8-ft and 32-ft walks) measuring gait speed, step length, cadence, and stride regularity; physical activity features (red) and gait quality features (green) extracted from daily-living accelerometer data. Classification performance, indicated by the area under the curve (AUC), improved from supervised-based features (AUC = 0.67 ± 0.09) to physical activity features (AUC = 0.69 ± 0.06), and was highest when using all available wrist features (AUC = 0.80 ± 0.06). A Friedman test showed a significant effect of feature set on AUC (p < 0.001). Post hoc Wilcoxon tests revealed the gait quality model outperformed both supervised (p < 0.001) and physical activity models (p < 0.01), with no significant difference between the latter two.

To statistically assess performance differences, we treated each outer cross-validation fold as an independent sample. A Friedman test revealed a significant overall effect of feature set on AUC (p < 0.001). Post hoc Wilcoxon signed-rank tests with a Bonferroni correction showed that the full daily-living gait model significantly outperformed the supervised assessment of gait model (p < 0.01) and also outperformed the PA model (p < 0.01). No significant difference was found between the supervised and PA models.

## DISCUSSION

The aim of this study was to extend and validate ElderNet for extracting spatial-temporal measures of gait from a wrist-worn accelerometer during daily living in older adults and individuals with gait impairments. Prior studies have reported that gait speed metrics derived from a wrist-worn accelerometer are associated with incident frailty and dementia^12,41^, yet the gait speed algorithms employed in prior studies have not previously been validated for use in older populations or those with gait impairments. Using real-world, daily-living wrist accelerometer recordings from a large cohort of well-characterized older adults and individuals with impaired gait, we optimized ElderNet to better support assessments in these groups. In addition, the current study includes preliminary clinical findings which demonstrate that multi-day mobility metrics extracted from a wrist-worn accelerometer can better discriminate adults with and without self-reported mobility disability. These metrics may improve subtyping of older adults compared to conventional gait performances and quantitative measures of total daily physical activity. Further work is needed to determine if different combinations of these metrics are differentially associated with disparate adverse health outcomes and can inform targeted therapies for impaired gait in aging adults.

For gait speed, ElderNet significantly outperformed the Soltani et al. state-of-the-art model^33^, and a supervised baseline model by achieving lower error rates alongside high R² and ICC on the Mobilise-D TVS test set. Moreover, it achieved superior performance on an external dataset of healthy young adults. The improved performance by ElderNet was most pronounced within the Mobilise-D TVS cohort, highlighting the benefits of optimizing ElderNet models for assessing older adults and populations with gait impairments. Additionally, ElderNet consistently demonstrated high reliability across all cohorts, with ICC(2,1) values exceeding 0.7— a threshold commonly used in state-of-the-art Mobilise-D studies^38^.

Fixed-placement 3D sensors on the lower back typically yield cleaner gait signals for multiday recordings compared to wrist-worn devices ^19^. However, ElderNet has been shown to significantly reduce this performance gap^20^. Indeed, ElderNet also demonstrated improved accuracy compared to a lower-back-based gait speed algorithm previously evaluated on the Mobilise-D cohort by Kirk et al.^23^. Furthermore, ElderNet achieved a lower mean error of 0.17 cm/s, compared to 6.00 cm/s reported in the prior study using the lower back sensor. It is important to note that Kirk et al. evaluated their algorithm using all available data without splitting into separate training and test sets, complicating a direct comparison with our approach, which was strictly evaluated on an independent test set. Furthermore, Kirk et al. only included walking bouts that were successfully detected by both their gait detection algorithm and the reference system. This selection criterion likely favored simpler and less variable walking bouts, potentially leading to overly optimistic performance. In contrast, our validation included all walking bouts identified by the reference system, likely encompassing a broader range of complex and challenging gait scenarios. Thus, despite methodological differences, our approach may provide a more robust and ecologically valid estimation of real-world gait performance.

Gait speed estimation performance of ElderNet remained robust across all cohorts within the Mobilise-D dataset, but was lowest in the MS cohort, which was much younger than the other Mobilise-D participants. Reduced performance in the younger MS cohort (Table 1) may be due to the fact that ElderNet was explicitly optimized for older adults during the SSL phase, having been pretrained on the RUSH MAP dataset, which is comprised exclusively of older individuals (Table 5). In the future, if more labeled data becomes available, it may be interesting to refine ElderNet to improve its performance in healthier and younger adults.

We further fine-tuned ElderNet to estimate additional gait quality metrics—cadence, stride length, and stride regularity—capturing diverse facets of mobility^42–44^. To evaluate the contribution of the SSL component, we compared ElderNet’s performance with that of a purely supervised baseline. For cadence and stride length, ElderNet consistently outperformed the supervised model across most evaluation metrics, with statistically significant improvements observed for all metrics except cadence R², which showed a positive trend. In contrast, no significant performance difference was observed for stride regularity. This may be due to the nature of the stride regularity labels, which were derived from lower-back accelerometer data, whereas the SSL phase of ElderNet relied solely on wrist-worn accelerometers. Incorporating multi-sensor data (e.g., gyroscopes, PPG^45^), including from additional body locations, into the SSL pre-training phase may enhance performance for such metrics. However, these enhancements would need to be balanced against increased system complexity, deployment costs, and the potential for reduced user compliance, and therefore warrant careful consideration in future research.

A potential advantage of deriving gait metrics from multi-day recordings is their ability to capture natural, within-subject variability in walking behavior, offering richer and more ecologically valid insights compared to traditional, short-duration supervised gait assessments. Here, we applied ElderNet on real-world data from the RUSH MAP, extracting meaningful statistics describing the distributions of the different gait quality metrics. Although gait metrics derived from supervised assessments were significantly associated with most of those obtained from daily living contexts, the observed correlations were weak-to-moderate (Fig. S1). This confirms previous findings reported using lower back-worn sensors that supervised and real-world gait measures, while related, likely reflect distinct aspects of gait and mobility^16–18^. The ability to extract accurate gait measures from a wrist-worn accelerometer in daily life, therefore, holds promise for extending these insights and improving diagnostic and prognostic capabilities.

Indeed, our analysis demonstrated that gait metrics derived from multi-day recordings provided superior discriminative power for mobility disability compared to conventional supervised assessments. Moreover, these real-world gait metrics also outperformed the model based on PA metrics derived from the wrist-worn accelerometer from which the gait metrics were derived. This highlights the added clinical value of extracting gait-specific measures from everyday living metrics, offering a more detailed and predictive assessment of mobility and health status.

It is worth noting that the clinical utility analyses in the current study were based on a basic machine learning approach, serving as a preliminary demonstration of clinical potential. Future research will aim to develop dedicated neural network-based classifiers specifically trained on clinical outcomes, integrating and potentially fine-tuning ElderNet directly on tasks such as mobility disability classification or the prediction of the development of adverse events such as mortality or the development of Alzheimer’s or Parkinson’s using multiday recordings of daily living. We anticipate that the application of more advanced deep learning techniques will further leverage the benefits of extracting gait measures from a wrist-worn sensor.

One limitation of this study is that ElderNet was validated on a test set drawn from the same distribution as the training dataset (i.e., the Mobilise-D dataset). In addition, the test set comprised a relatively small sample size (n=18), which may affect the robustness of our conclusions. To address these limitations, we employed an external dataset of healthy young adults (n=11) and further utilized the RUSH MAP dataset (n=423) to extend the validation. Future studies should investigate ElderNet’s effectiveness on additional external labeled datasets to further enhance its generalizability and clinical utility.

Recent work with UK Biobank accelerometry data highlights the potential feasibility of using daily living information to stratify older adults at risk for Parkinson’s disease^46,47^. However, this study did not exploit detailed multi-day gait-specific measures and suffered from other limitations. By integrating spatial-temporal gait metrics derived from multi-day recordings, there is an opportunity to significantly enhance the early detection and characterization of such conditions.

Overall, our findings demonstrate that ElderNet, previously validated for accurately detecting walking bouts from wrist-worn accelerometers, can now reliably estimate spatial-temporal gait metrics during everyday life. This capability has been specifically validated in older adults with age-related gait impairments and neurological conditions. Utilizing wrist-worn accelerometers for remote phenotyping offers substantial practical advantages, including affordability, widespread acceptance by older adults, high compliance, and suitability for long-term unobtrusive monitoring. Historically, detailed gait characterization has been challenging due to the complexity of wrist-worn accelerometer signals—a limitation directly addressed by our approach. By overcoming these barriers, ElderNet advances aging research, enabling the identification of specific gait alterations that may serve as biomarkers for diseases or functional impairments. Ultimately, this could facilitate the development of personalized interventions informed by an individual’s unique mobility patterns.

## METHODS

### Study Design

This study consisted of three primary stages:

1. **Estimation of Spatial-Temporal Gait measures**: Extending ElderNet to quantify continuous gait quality metrics.
2. **Model Validation and Comparison**: Comparing ElderNet’s performance against state-of-the-art and baseline supervised models using internal and external datasets.
3. **Demonstration of Potential Clinical Utility**: Evaluating the clinical relevance of gait metrics by applying ElderNet to real-world daily living data from cohorts with varying clinical characteristics.

### Datasets and Participants

Four independent datasets were used across different phases of model development, validation, and evaluation of clinical utility:

#### Dataset 1: UK Biobank

a large-scale dataset comprising one week of wrist-worn accelerometry recordings from over 100,000 individuals (mean age 62.1 ± 7.9 years). The UK Biobank dataset was used indirectly via the publicly available ResNet deep learning model pre-trained using SSL by Yuan et al.^25^. This model served as the fixed feature extraction backbone for ElderNet.

#### Dataset 2: Rush Memory and Aging Project

(MAP), which includes 819 older adults (mean age 83.4 ± 7.3 years) who wore a wrist accelerometer for up to 10 consecutive days. This dataset was used in two stages: (i) to adapt ElderNet to the mobility patterns of older adults through additional SSL pretraining, and (ii) to evaluate the clinical utility of ElderNet by classifying mobility disability based on multi-day gait metrics. A subset of 423 individuals (mean age 83.6 ± 6.9 years) also underwent standardized supervised gait assessments, enabling comparison between real-world and clinical gait measures.

#### Dataset 3: Mobilise-D Technical Validation Study

This multicenter observational study comprised 108 participants recruited from five clinical centers across the UK, Germany, and Israel. Following the exclusion of 23 individuals due to incomplete recordings, 85 participants remained for analysis. The cohort included both healthy older adults and individuals diagnosed with one of five conditions: chronic obstructive pulmonary disease (COPD), Parkinson’s disease (PD), multiple sclerosis (MS), proximal femoral fracture (PFF), and congestive heart failure (CHF). Each participant wore an inertial unit including a tri-axial accelerometer on the non-dominant wrist, a lower-back inertial unit, and the INDIP multi-sensor reference system for up to 2.5 hours during their typical daily activities (at home, work, in the community, or outdoors). Participants were also encouraged to perform a limited set of predefined tasks (outdoor walking, walking up and down a slope and stairs, and moving from one room to another) ^48^. The INDIP system, which integrates complementary sensing modalities—including two plantar pressure insoles, three inertial units, and two distance sensors—was used to detect walking bouts and provide ground-truth annotations of spatio-temporal gait metrics at the stride level^39,40^.

This dataset was used for hyperparameter tuning, training, and evaluation of ElderNet. It was partitioned into training (70%), validation (10%), and test (20%, n=18) sets, with subject-wise partitioning to prevent data leakage. All three sets—training, validation, and test—include representation from all clinical cohorts in the Mobilise-D study (i.e., HA, CHF, COPD, MS, PD, and PFF), ensuring balanced evaluation across the diverse participant population. All performance comparisons—including against baseline supervised and state-of-the-art models—were conducted on the held-out test set. Ethical approvals for data collection were obtained from the relevant institutional review boards at all participating sites. A complete description of the data collection protocol is available in Mazza et al.^38^.

#### Dataset 4: Mobilise-D Healthy Young Adults (External Validation)

An independent valida-tion dataset comprising 11 healthy young adults (mean age 29.6 ± 7.8 years), collected using the same protocol as the Mobilise-D TVS. These data were used to assess ElderNet’s generalizability in gait speed estimation to a younger, healthier, and demographically distinct population.

### Data Preprocessing

The INDIP reference system was first used to identify the exact start- and end-times of every walking bout within the approximately 2.5-hour recording^19^. The corresponding wrist-accelerometer segments were extracted, ensuring that non-walking periods were discarded. Each consecutive bout lasting longer than 10 seconds was then partitioned into 10-second windows with 90 % overlap (i.e., a stride of 1 second), where windows never spanned two different bouts. We chose 10-second windows to align with the pre-trained UK Biobank model^25^ and added 90% overlap to enhance the temporal resolution of our analysis. All extracted segments were resampled to 30 Hz, matching ElderNet’s pre-training data from UK Biobank^25^. Because typical gait frequencies are below 10 Hz, this rate satisfies the Nyquist criterion and preserves all relevant signal content.

### Spatial-Temporal Gait Metrics

For every 10-second walking window, we derived four reference outcomes from the INDIP system: gait speed (cm/s), stride length (cm), cadence (steps/min), and stride regularity (arbitrary units, on a scale of 0 to 1). Speed and stride length were defined as the median of all complete INDIP-detected strides whose start and end both fell inside the window. Cadence was obtained by dividing the number of strides by the 10-second window duration and multiplying by six (one stride equals two steps). Stride regularity was calculated following Moe-Nilssen and Helbostad^49^. Briefly, the vertical acceleration of the lower-back device was demeaned and normalized, an unbiased autocorrelation function was applied to the 10-second segment, and the coefficient at a time lag corresponding to the mean stride duration (the second dominant peak) was taken as the stride-regularity index (range 0–1, with 1 denoting perfectly periodic strides). Each analysis window, therefore, carried a fully time-aligned ground-truth label for all four gait outcomes.

### ElderNet Model Development and Training

ElderNet, originally developed as a classification model for gait detection^20^, was adapted for the regression-based estimation of continuous gait metrics through a two-stage process. In the first stage, we leveraged a ResNet model that was pre-trained using SSL on the large-scale UK Biobank wrist-worn accelerometry dataset^5,25^. This pre-trained backbone served as a fixed feature extractor, with all ResNet weights frozen. To adapt the model for older adults, we added three fully connected layers on top of the frozen backbone and trained these layers using SSL in a multi-task learning framework on data from the RUSH MAP, which includes wrist-worn accelerometer recordings of older populations. The output of this stage was a 128-dimensional feature vector for each 10-second accelerometry window.

In the second stage, this RUSH-tailored SSL model was repurposed as a feature extractor for gait quality estimation. To enable regression outputs, we extended the model with 0–3 additional layers (selected via hyperparameter tuning) that mapped the 128-dimensional feature representation to continuous gait metrics such as gait speed. During this fine-tuning stage, the entire model, including the ResNet backbone and the newly added layers, was trained end-to-end using the training data from the Mobilise-D dataset (Dataset 3). This approach allowed ElderNet to learn precise, task-specific mappings while benefiting from the robust representations learned during the SSL phase.

Because stride length can also be expressed as walking speed divided by cadence, we computed two independent stride-length estimates for every 10-second window: one taken directly from the ElderNet output trained on stride-length labels, and a second obtained by dividing ElderNet’s predicted speed by its predicted cadence. We then reported the simple average of these two values as the final stride-length prediction, an approach that consistently reduced error compared with relying on either estimate alone.

Hyperparameter optimization was performed on the training (70% of dataset 3) and validation (10% of dataset 3) sets using the Optuna Python library with 50 trials^50^, employing five-fold cross-validation stratified by clinical cohort and grouped by participants. The hyperparameter search space is detailed in Supplementary Table S2. Model training incorporated early stopping with a patience of five epochs, selecting the configuration yielding the lowest MAE on the validation set. The loss function used was L1.

The model was implemented using PyTorch in Python 3.10 and trained using an NVIDIA RTX A4000 GPU with 16GB RAM. The training dataset consisted of 49,372 windows of 10 seconds each. During the final training with the optimal hyperparameter configuration, the average processing throughput was approximately 2,541.15 windows per second. For inference, the test set contained 13,998 windows, with a total inference time recorded at 8.55 seconds.

### Model Evaluation and Comparison

ElderNet’s performance was evaluated against a fully supervised baseline model with an identical ResNet architecture (without SSL pre-training) to quantify the benefit derived from the SSL component. Additionally, for gait speed, we compared ElderNet to the non-personalized biomechanically-based approach proposed by Soltani et al.^33^. This approach uses regression-based modeling with predefined biomechanical features extracted from wrist-worn accelerometry data, including cadence, acceleration energy, mean absolute jerk, hand-swing intensity, mean acceleration norm, and altitude change of path. Given the absence of barometric data in our dataset, altitude-related features were excluded from the model. We trained Soltani’s regression model using the same training dataset as ElderNet (i.e., 80% of dataset 3) and evaluated it on the identical test dataset (i.e., 20% of dataset 3, and external dataset 4) to ensure a fair and consistent comparison. We considered the method proposed by Soltani et al. as a state-of-the-art benchmark due to its strong real-world performance in gait speed estimation, achieving a median RMSE of approximately 10 cm/s and demonstrating significant associations with clinical metrics such as frailty and handgrip strength^41^.

### Demonstration of Potential Clinical Utility

To evaluate the clinical utility of gait metrics derived from ElderNet, we applied the fine-tuned models to real-world wrist accelerometry data collected during daily life from 819 community-dwelling older adults participating in the RUSH MAP^36,37^ (Dataset 2). Among these participants, 423 underwent additional supervised gait assessments using lower-back accelerometry during standardized 8-foot and 32-foot walking tests. For each of the supervised walking tests, we extracted four measures corresponding to the daily living gait quality metrics: gait speed, stride length, cadence, and stride regularity^51,52^.

Our aim was to determine whether multi-day gait metrics could capture aspects of mobility that are not fully represented by brief supervised assessments or general physical activity measures. Daily-living walking bouts were identified from the wrist-worn accelerometry recordings using the previously validated gait detection algorithm implemented in ElderNet^20^. From each identified walking bout, we extracted four gait quality metrics—gait speed, stride length, cadence, and stride regularity—using the fine-tuned ElderNet regression models. Additionally, stride regularity was independently calculated directly from the raw wrist accelerometry data using an autocorrelation-based signal processing method analogous to the approach applied to lower-back data^49^.

Participants with fewer than three valid days of wrist accelerometry data were excluded, resulting in a final sample of 787 participants^8^. For each participant, comprehensive summary statistics—including mean, median, standard deviation, kurtosis, skewness, range, and decile percentiles—were calculated at both the 10-second window level and across aggregated walking bouts. In parallel, physical activity measures were derived from the magnitude of the same 3D acceleration signal, summarized over a daily time frame. These daily summaries included the same set of statistics used for gait quality metrics and were computed for both the entire recordings and the segments identified as walking bouts. A detailed overview of all extracted features is provided in Table S3.

Finally, to assess the clinical relevance of the derived gait metrics, we constructed classification models using an XGBoost classifier (implemented via the scikit-learn library) to discriminate between participants with and without mobility disability, as defined by the Rosow-Breslau scale (dichotomized into no disability versus disability)^53^. Three separate classification models were implemented: one based on gait features from supervised assessments, a second using physical activity features from daily living recordings, and a third incorporating daily living gait metrics. Model performance was evaluated using nested cross-validation with 5-fold inner and 10-fold outer loops with subject-wise partitioning to prevent data leakage. Hyperparameter tuning was performed via a randomized search (see Table S4 for the hyperparameter ranges). Model evaluation was based on Receiver Operating Characteristic (ROC) analysis, with the AUC serving as the primary metric.

### Statistical Analysis

Statistical analyses for the estimation of gait metrics (Datasets 3 and 4) were conducted at the subject level: model-performance metrics were first aggregated over the repeated 10-s windows contributed by each participant, so every statistical test compares independent subjects. Shapiro–Wilk tests rejected normality; therefore, non-parametric, two-sided tests were used throughout.

For gait speed, where three models (ElderNet, a supervised baseline, and a state-of-the-art comparator) were compared, we used the Friedman test to assess overall differences across models. If the Friedman test indicated statistical significance, post hoc pairwise comparisons were conducted using the two-sided Wilcoxon signed-rank test, with Bonferroni correction applied to account for multiple comparisons. For the other gait metrics (cadence, stride length, and stride regularity), comparisons involved only two models (ElderNet and its supervised counterpart). Therefore, we used the Wilcoxon signed-rank test to evaluate differences in subject-level performance, with p-values adjusted for multiple comparisons via the Bonferroni method.

For the classification task in the RUSH MAP (Dataset 2), the AUC from each of the ten outer cross-validation folds was treated as an independent observation. To compare the three competing feature sets (supervised gait metrics, PA metrics, and daily-living gait metrics), we employed the Friedman test on fold-level AUCs. When the Friedman test revealed significant differences, we conducted post hoc two-sided Wilcoxon signed-rank tests with Bonferroni correction for pairwise comparisons.

All statistical analyses were performed in Python using the scipy.stats library. Statistical sig-nificance was set at p < 0.05 after correction for multiple comparisons.

## Data Availability

The dataset from the Mobilise-D technical validation study (Dataset 3) can be found on Zenodo: https://doi.org/10.5281/zenodo.13899386. All other data and related algorithms (i.e., Dataset 2) included in these analyses are available via the Rush Alzheimer's Disease Center Research Resource Sharing Hub, which can be found at www.radc.rush.edu. It has descriptions of the studies and available data. Any qualified investigator can create an account and submit requests for de-identified data.

https://www.radc.rush.edu/

## Code availability

The code will be made publicly available on GitHub upon publication.

## ACKNOWLEDGMENTS

The authors gratefully acknowledge the entire Mobilise-D Work Package 2 team for their ongoing collaboration, insightful discussions, and valuable contributions. Special thanks are extended to the study participants for their time, commitment, and enthusiasm, especially during the challenging circumstances of the COVID-19 pandemic. The authors also sincerely thank the contributors to the RUSH Memory and Aging Project, as well as the dedicated staff at the Rush Alzheimer’s Disease Center for their continuous support. Y.E.B. received research support from the Aufzien Academic Center and the Herczeg Institute on Aging at Tel-Aviv University.

This work was supported in part by grants from the NIH (R01AG017917; R01AG056352, R01AG79133, R01AG078256) and by the Mobilise-D project. The Mobilise-D project received funding from the Innovative Medicines Initiative 2 Joint Undertaking (JU) under grant agreement No. 820820. This JU receives support from the European Union’s Horizon 2020 research and innovation program and the European Federation of Pharmaceutical Industries and Associations (EFPIA). SDD, AJY, and LR were supported by the IDEA-FAST project that has received funding from the Innovative Medicines Initiative 2 Joint Undertaking (JU) under grant agreement No. 853981. SDD, AJY, and LR were supported by the National Institute for Health Research (NIHR) Newcastle Biomedical Research Centre (BRC) based at The Newcastle upon Tyne Hospital NHS Foundation Trust, Newcastle University, and the Cumbria, Northumberland and Tyne and Wear (CNTW) NHS Foundation Trust. SDD, AJY, and LR were also supported by the NIHR/Wellcome Trust Clinical Research Facility (CRF) infrastructure at Newcastle upon Tyne Hospitals NHS Foundation Trust. SDD was supported by the UK Research and Innovation (UKRI) Engineering and Physical Sciences Research Council (EPSRC) (Grant Ref: EP/X031012/1 and Grant Ref: EP/X036146/1).

All opinions are those of the authors and not the funders. The content in this publication reflects the authors’ view, and neither IMI nor the European Union, EFPIA, NHS, NIHR, or any associated partners are responsible for any use that may be made of the information contained herein.

## AUTHOR CONTRIBUTIONS

Participant recruitment and clinical oversight: W.M., C.B, J.M.H., A.J.Y., L.R. Algorithm development: Y.E.B., O.P., J.M.H. Data analysis, statistical analysis: Y.E.B. Figures and tables preparation: Y.E.B. Data interpretation: Y.E.B., J.M.H., O.P. Drafting of the initial manuscript: Y.E.B., J.M.H., O .P. Intellectual contribution: Y.E.B., F.K., L.P., C.B., A.C., W.M., B.V., A.J.Y., L.R., S.DD., A.M., A.S.B., J.M.H., O.P. All authors have provided critical intellectual input during the revision of the manuscript. All authors have reviewed the manuscript and approved the submitted version.

## ETHICS DECLARATION

### Ethics approval and consent to participate

For the Rush Memory and Aging project, all participants provided written informed consent before participation. The study was approved by the Rush University Medical Center Institutional Review Board and conducted in accordance with the Declaration of Helsinki. For the Mobilise-D study, ethical approval was obtained at the individual sites (London-Bloomsbury Research Ethics Committee, 19/LO/1507; Helsinki Committee, Tel Aviv Sourasky Medical Center, Tel Aviv, Israel, 0551-19TLV; ethical committee of the medical faculty of The University of Tübingen, 647/2019BO2; ethical committee of the medical faculty of Kiel University, D438/18) and all participants gave informed consent before participating.

