## Supplementary material for "Continuous Assessment of Daily-Living Gait Using Self-Supervised Learning of Wrist-Worn Accelerometer Data"

**Table S1**. Comparison of ElderNet performance with state-of-the-art and supervised models for gait speed estimation across different test sets.

| **Model** | **Test Set** | **MAE (cm/s)** | **RMSE**  **(cm/s)** | **MAPE (%)** | $\boldsymbol{R}^{\boldsymbol{2}}$ | **ICC** |
| --- | --- | --- | --- | --- | --- | --- |
| **ElderNet** | **Dataset 3** | 8.8  [6.53 11.02] | 10.7  [8.59 13.75] | 12.73  [9.45 15.58] | 0.74  [0.47 0.80] | 0.87  [0.78 0.92] |
|  | **Dataset 4** | 8.3  [6.53 11.79] | 11.4  [8.85 14.49] | 8.22  [6.66 11.04] | 0.72  [0.57 0.81] | 0.87  [0.82 0.92] |
| **Soltani et al.** | **Dataset 3** | 16.3  [12.12 18.17] | 19.0  [13.99 22.30] | 21.27  [17.19 26.70] | 0.20  [-0.8 0.6] | 0.62  [0.43 0.78] |
|  | **Dataset 4** | 12.9  [9.73 15.13] | 14.8  [12.89 17.67] | 11.39  [8.86 15.23] | 0.68  [0.42 0.73] | 0.84  [0.72 0.87] |
| **Supervised** | **Dataset 3** | 14.1  [11.33 18.06] | 17.4  [14.92 22.21] | 21.63  [16.49 31.11] | 0.23  [-0.22 0.58] | 0.59  [0.38 0.72] |
|  | **Dataset 4** | 13.8  [9.58 16.51] | 17.9  [12.91 19.80] | 15.85  [9.38 17.23] | 0.53  [0.38 0.65] | 0.74  [0.68 0.81] |

Performance is reported at the subject level as the median (with the 25th and 75th percentiles in brackets). The internal test set (Dataset 3) was the Mobilise-D TVS set, which included 18 participants (20% of the dataset). The external test set (Dataset 4) included 11 healthy young adults.

Table S2. Hyperparameter search space used for ElderNet fine-tuning in gait quality estimation.

| **Hyperparameter** | **Values** |
| --- | --- |
| Learning rate | 1e-5, 5e-5, 1e-4, 5e-4, 1e-3 |
| Batch size | 64, 128, 256, 512, 1024 |
| Number of layers (regression head) | 0-3 |
| Batch norm (for regression layers) | Yes/No |
| Weight Decay | 0, 0.01, 0.1 |

Table S3. Summary of gait quality and physical activity features.

| **Measure** | **Extraction Level*** | **Unit** | **Summary Statistics Computed** | **Distribution Features**** |
| --- | --- | --- | --- | --- |
| **Gait Measures** |  |  |  |  |
| Gait Speed | 10-sec window, Walking bout | cm/s | Median, Mean, Std, Percentiles (10th–90th), Kurtosis, Skewness, Range | Histogram bins (10 bins across 0–1.8 cm/s) |
| Stride Length | 10-sec window, Walking bout | cm | As above | Histogram bins (10 bins across 0–2 cm/stride) |
| Cadence | 10-sec window, Walking bout | Steps per minute | As above | Histogram bins (10 bins across 40–160 steps/min) |
| Stride Regularity (Model-based) | 10-sec window, Walking bout, | A.U (0-1) | As above | Histogram bins (10 bins across 0–1); |
| Stride Regularity (Signal Processing) | 10-sec window, Walking bout | A.U (0-1) | As above | Histogram bins (10 bins across 0–1) |
| **Physical Activity Measures** |  |  |  |  |
| Average magnitude of the acceleration signal | Day-level and Walking bout level | m/s² | Median, Mean, Std, Percentiles, Kurtosis, Skewness, Range | — |
| Standard deviation of the acceleration magnitude signal | Day-level and Walking bout level | m/s² | As above | — |
| Minimum of the acceleration magnitude signal | Day-level and Walking bout level | m/s² | As above | — |
| Maximum of the acceleration magnitude signal | Day-level and Walking bout level | m/s² | As above | — |

***Extraction Level**:

1. 10-second window level: Statistics were computed over overlapping windows (10-second windows, with 9-second overlap).
2. Walking bout level: Windows were aggregated over contiguous bouts of walking activity.
3. Day-level: Aggregated features computed over the entire day.

****Distribution Features**:

The global ranges used to compute the histogram-based distribution features were determined by combining empirical observations from our dataset with normative values reported in the literature^1,2^. For each gait measure, the observed values were divided into ten equal-width bins across a predefined range. For example, gait speed was set between 0 and 1.8 m/s—reflecting the upper limits of walking speeds typically observed in older adults—while cadence was bounded between 40 and 160 steps per minute. Stride length was assigned a range of 0 to 2 m (i.e., 0–200 cm), and both the model-based and signal processing–based stride regularity metrics were normalized between 0 and 1. Each bin feature represents the number of 10-second windows (or aggregated bouts) during which the participant’s measure falls within that interval (e.g., the bin covering 0.6–0.8 m/s indicates the count of windows with gait speeds in that range).

**Table S4.** Hyperparameter Ranges for the XGBoost Classifier

| **Hyperparameter** | **Values** |
| --- | --- |
| n_estimator | 10, 50, 150, 300, 500 |
| learning_rate | 0.001, 0.005, 0.01, 0.05, 0.1, 02 |
| subsample | 0.1, 0.3, 0.6, 0.9 |
| colsample_bytree | 0.5, 0.7, 0.8, 0.9 |
| gamma | 0, 0.1, 0.2, 0.3, 0,5 |
| alpha | 0, 0.01, 0.1, 1 |
| lambda | 1, 2, 3 |

All hyperparameters follow the sklearn-compatible API of xgboost.XGBClassifier.
Hyperparameter optimization was conducted using RandomizedSearchCV from scikit-learn with 100 iterations.

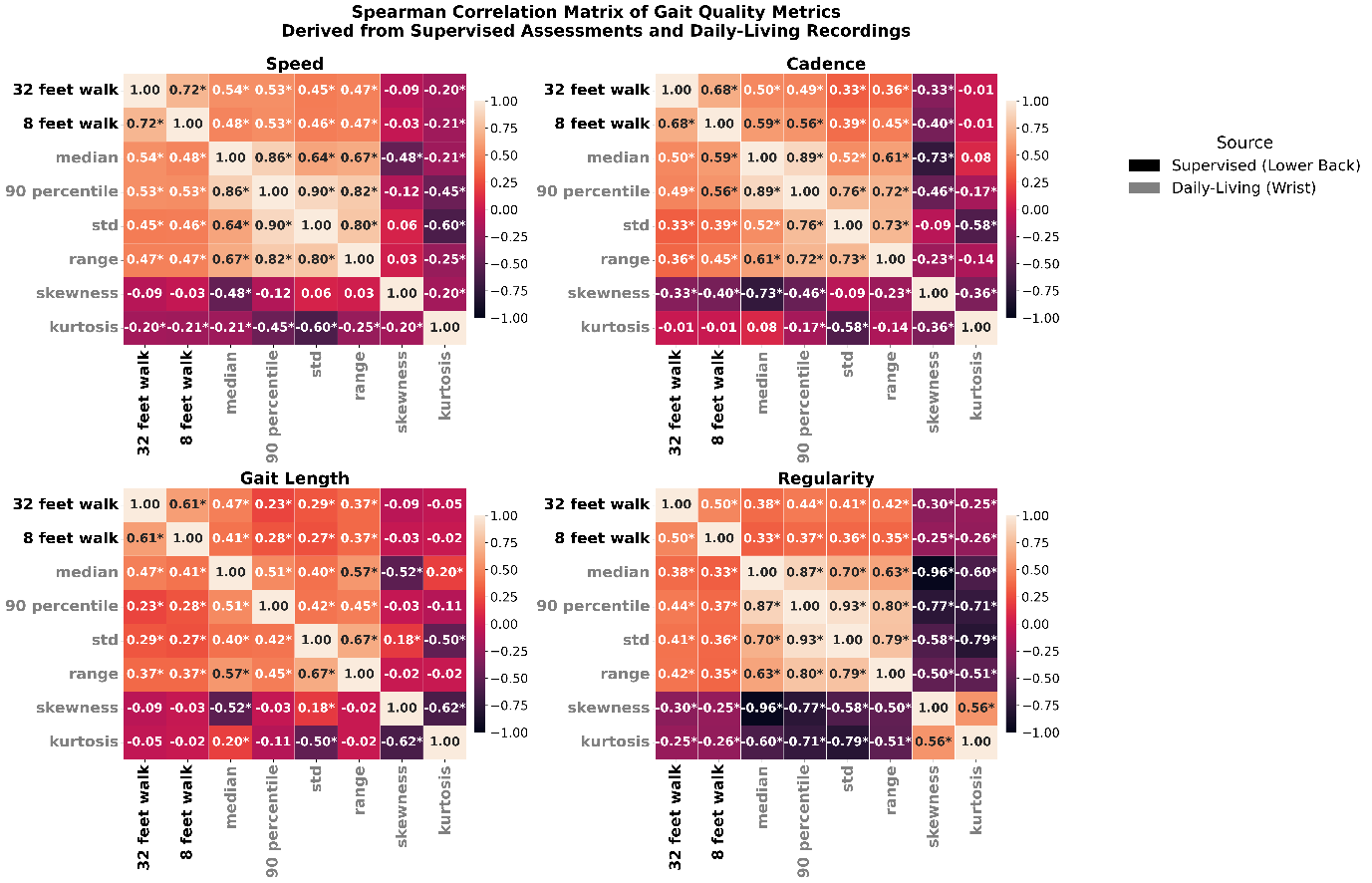

**Figure S1**. Pairwise correlations between gait measures of speed, cadence, gait length, and regularity obtained from supervised assessments (black labels: 32 feet walk, 8 feet walk) and daily living recording (grey labels: “median,” “90 percentile”, “std,” “range,” “skewness”). Each cell shows the Spearman correlation coefficient of the two variables. Diagonal cells are self-correlations (1.00). An asterisk (“*”) indicates statistical significance (p < 0.05) after Bonferroni correction. While some daily-living metrics correlate with supervised assessments, much of the variance remains unexplained, suggesting that wrist-derived measures reflect supervised performance but also capture additional real-world gait characteristics.
